# ‘We need this now’: Acceptability and usability of a pediatric antiretroviral microarray patch prototype in South Africa and Uganda

**DOI:** 10.64898/2026.09.17.26363333

**Authors:** Manjari Quintanar-Solares, Justine Komunyena Tumusiime, Pinky Mbotho, Nolwazi Ngcobo, Priscilla Kwarteng, Thobekile Sibaya, Jessica Joyce Mistilis, Moherndran Archary, Robert K.M. Choy

## Abstract

Children living with HIV face significant challenges with adherence to daily oral antiretroviral (ARV) treatment due to poor palatability, complicated dosing, and caregiver burden. Long-acting delivery systems designed specifically for children may help overcome these barriers. Microarray patches, also known as microneedle patches, are a needle-free drug delivery technology that could simplify treatment administration and improve adherence. This study explored the acceptability and usability of a pediatric ARV microarray patch prototype among stakeholders in South Africa and Uganda, two countries with a high burden of pediatric HIV.

A formative study was conducted in 2024 with 48 participants, including caregivers, community health workers, health care providers, and decision-makers. Data were collected through qualitative methods: interviews, simulated use sessions, and observations. Participants demonstrated high usability of the prototype, with 97% completing all application steps and nearly all reporting the prototype as easy to use and expressing confidence in successful administration. Most participants found the prototype’s size, wear time, and temporary skin changes acceptable, and all preferred a weekly patch over daily oral treatment. Applying multiple patches at once to achieve a therapeutic dose was considered feasible. Decision-makers foresaw strong programmatic fit of a pediatric ARV microarray patch if caregivers apply the patch at home following initial facility-based guidance.

Overall, the pediatric ARV microarray patch prototype was perceived as highly acceptable and usable across diverse end users, findings that generally align with studies of microarray patches for other use cases (e.g., contraception, HIV pre-exposure prophylaxis, vaccine delivery). The simplicity, comfort, and convenience of a pediatric ARV microarray patch could substantially improve treatment adherence and quality of life for children living with HIV. Further development and clinical evaluation are warranted to advance this promising technology toward real-world use.

## Introduction

Globally, in 2024, an estimated 1.4 million children (0–14 years) were living with HIV, but only 55% received any treatment and 75,000 of these children died [1]. The HIV burden is highest in sub-Saharan Africa, accounting for approximately 86% of all children living with HIV [2]. At the end of 2022, approximately 230,000 children 0 to 14 years of age in South Africa were living with HIV, and about 80,000 children 0 to 14 years of age in Uganda were living with HIV [3,4]. While HIV treatment of children across eastern and southern Africa has improved in recent years to an average of 65% coverage in 2023, an average of only 54% of children on antiretroviral (ARV) therapy in these areas were virally suppressed [5]. Globally, in the same year, treatment coverage among those aged 15 years and over was an average of 84%, considerably higher than among children.

The challenges affecting adherence to pediatric HIV treatment have been well documented by researchers and clinicians, and include the lack of appropriate child-friendly formulations [6,7,8,9,10]; onerous or complicated dosing [11,12,13,14], such as mixing of several drugs, multiple doses per day, or frequent refills; and poor palatability [12,15,16,17], causing refusal, spitting, or vomiting of the medication. These challenges, among others, lead to decreased adherence, which in turn contributes to failure to suppress viral replication completely and thus increased morbidity and mortality.

A long-acting drug delivery system that is designed to meet the unique needs of pediatric HIV patients could improve access and adherence to ARV treatment [18]. One such platform is microarray patches (MAPs). MAPs, also known as microneedle patches, are a novel, needle-free delivery technology in development for some drugs and vaccines. MAPs are applied like an adhesive bandage and contain an array of micron-scale projections that pierce the stratum corneum and facilitate delivery into the skin (Fig. 1).

**Fig. 1.**
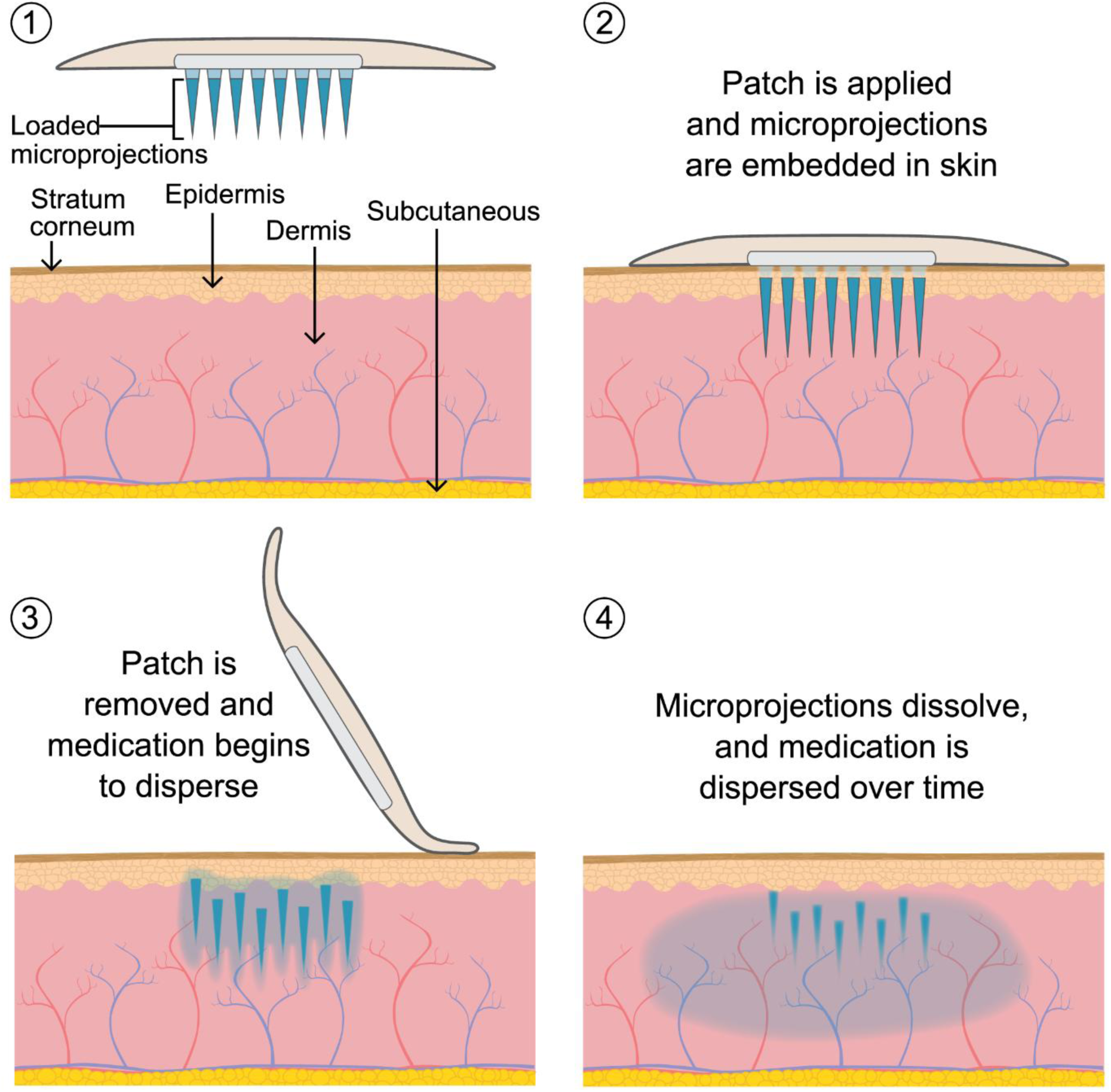
Microarray patch delivery.

Due to their ease of use and expected acceptability (they are less painful than injections) [19,20,21], MAPs can potentially improve adherence to treatment, particularly when compared to unpalatable or complicated oral options for children. The Pediatric Antiretroviral Drug Optimization group (known as PADO), led by the World Health Organization, considers MAPs to be one of the most promising technologies for HIV treatment and prevention for children, particularly for children younger than 2 years [22].

PATH developed a low-cost, easy-to-use MAP indicator prototype (Fig. 2) with the following target characteristics, based on prior feedback received from global and country stakeholders: similar size to a postage stamp, to be applied in the upper back to avoid soiling and removal by children, and using multiple MAPs at once to achieve required dose for different weight bands. The indicator is attached to the baseplate of the microneedle array. Its purpose is to confirm that enough pressure has been applied to insert the micron-scale projections into the skin. Once applied, the indicator can be removed. An adhesive holds the array of microneedles on the skin for the required wear time until the microneedles separate from the baseplate, thereby releasing the drug into the skin. The adhesive can then be removed and discarded. The MAP indicator prototype was round in shape with a 3-cm diameter, similar in size to a postage stamp, and it included three cues (visual, auditory, and tactile) that would enable users to confirm successful application.

**Fig. 2.**
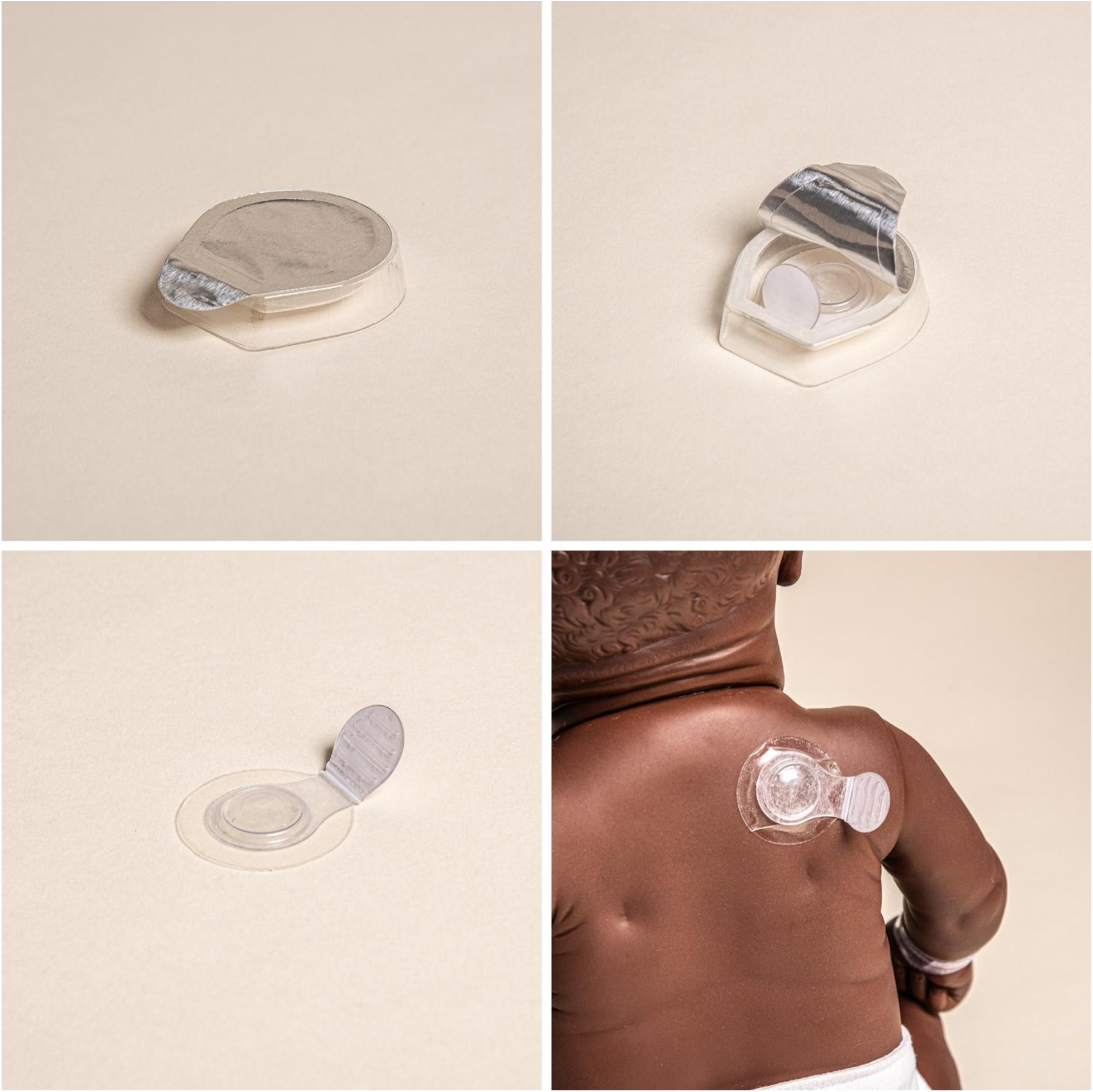
Microarray patch indicator prototype for pediatric antiretroviral drugs.

PATH and other partners have also been conducting pharmacokinetic modeling and animal testing for three promising ARV drugs [23,24]. The preliminary results of this modeling allowed PATH to calculate how much drug could be loaded to each array, considering the acceptable size, and thus how many MAPs would need to be applied at once to achieve the needed dose to deliver a therapeutic effect for each of the World Health Organization ARV weight bands [25,26]. Preliminary data suggest the feasibility of a weekly or monthly schedule, depending on the drug’s potency, and the size and number of arrays.

This study aimed to explore the use case and programmatic fit of a pediatric ARV MAP with stakeholders in two high HIV burden countries, and to assess the acceptability and usability of the PATH MAP indicator prototype with end users in these countries. Results from the study will be used to refine the prototype design, set critical attributes for the formulation of the arrays, and advance development of a pediatric ARV MAP toward clinical testing.

## Methods

### Study design and setting

We conducted a formative study that utilized qualitative methods: in-depth interviews, simulated use, and user observations. The study was conducted in two countries with a high burden of pediatric HIV: South Africa and Uganda. Participants were recruited between April 23 and September 27, 2024, from urban and rural areas, and included country-level decision-makers and target end users. Sampling was purposive to obtain a variety of opinions. In South Africa, participants were recruited at the national level and from eThekwini and uMkhanyakude Municipalities. In Uganda, participants were recruited at the national level and from Fort Portal, Kampala, Masaka, and Mbale Districts. Potential participants were identified by their role at the national level or by their affiliation with a regional HIV care center, where children receive the standard of care (oral ARV treatment) according to their respective country’s ARV treatment guidelines. Country-level decision-makers were approached by phone to gauge their interest in participating. Health care providers were suggested by the leadership of each regional HIV care center and then approached in person by a study representative. Community health workers and caregivers were suggested by health care providers and then approached in person.

The country-level decision-makers interviewed were experts in pediatric HIV programs and care. They participated in an in-depth interview and were asked about the acceptability of the ARV MAP and its attributes, foreseen use cases, and other programmatic considerations.

Target end users of the pediatric ARV MAP were those who provide direct care to children living with HIV: health care providers, community health workers, and caregivers. These target end users were engaged in simulated use, observation, and an in-depth interview in their preferred local language. During simulated use, study staff introduced MAPs to participants, following a script (S1 Appendix). Data collectors explained how MAPs work by showing participants the drawings from the script and using simple language (“patch with fast-dissolving microprojections penetrate the skin, then the backing is removed and the medication disperses over time”) and explained the target characteristics of the pediatric ARV MAP. To avoid confusion and the fear often associated with hypodermic metal needles, the term “microneedles” was intentionally *not* used. Participants were then trained on the administration of the MAP indicator prototype on an infant manikin and were provided with a job aid and an opportunity to practice handling and applying the prototype. Participants were then asked to administer the prototype on the infant manikin on their own, guided only by the job aid, while the data collector observed the simulation. Afterward, participants were interviewed on their use experience and acceptability of the ARV MAP and its attributes and were asked for additional input. Caregivers of different ages, who care for children of different age groups, were recruited to ensure various levels of visual acuity and dexterity and a variety of ARV administration experiences.

Interviews with country-level decision makers were conducted in a private space, either in person inside a health facility, or virtually during a video call. Study activities with target end users (interviews, simulated use, and observation) were conducted in person, in a private space inside a health facility or a research office. In South Africa, engagement with participants was conducted in English and isiZulu. In Uganda, engagement with HIV experts and health care providers was conducted in English, and engagement with community health workers and caregivers was conducted in Luganda, Lugisu, Rutooro, and English.

For perceived usability, simulation participants were asked to answer using the following Likert-type scales: (1) ease of use (very easy, easy, somewhat easy, somewhat difficult, difficult, very difficult); (2) confidence in administering the prototype successfully (very confident, confident, fairly confident, a little bit confident, not at all confident).

All participants were also asked about their preference for pediatric ARV treatment delivery modality, comparing MAPs to existing options (i.e., daily oral treatment) and pediatric long-acting treatment options currently in development and undergoing clinical trials [27,28,29], such as intramuscular or subcutaneous injections administered every 2 to 6 months.

Data collectors were fluent in local languages and received training in obtaining informed consent for human research subjects, use of the prototype, use of the data collection tools (interview and observation guide), and qualitative interviewing methodology (e.g., probing style, avoiding leading statements). Notes were taken by data collectors during all data collections activities. Interviews were audio recorded and used to complete notes taken.

### Materials

The MAP indicator prototype included primary packaging and the indicator itself, with medical-grade adhesive, but contained no drugs or micron-scale projections (Fig. 3). The three indicator cues for successful application were *tactile* (feeling the dome invert), *auditory* (hearing a snap upon pressing the dome), and *visual* (seeing the dome change color from transparent to white). The prototypes used in this study were manufactured in PATH’s product development engineering laboratory.

**Fig. 3.**
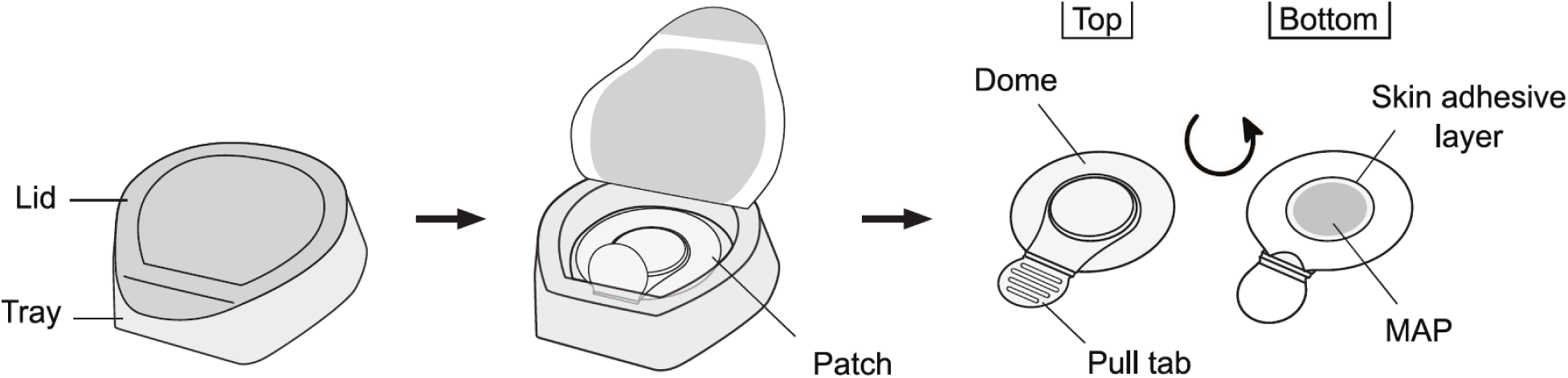
Parts of the PATH microarray patch indicator prototype.

A one-page job aid (in the preferred local language) illustrated the steps for using the indicator prototype (S2 Appendix). A newborn-sized, nonmedical baby care model (3B Scientific; Tucker, Georgia, USA) was used to simulate an infant, and Tegaderm™ (3M, Maplewood, Minnesota, USA) was applied on the upper back of the baby care model, prior to simulation, to provide a surface onto which the prototype could adhere in a manner that more accurately mimicked human skin than the surface of the manikin.

### Analysis

Frequencies for usability metrics (observed and participant reported) and acceptability metrics (participant reported) were calculated as percentages. These counts were used descriptively to illustrate the relative prominence of themes rather than for statistical inference. Participant input on programmatic fit, acceptability and preference rationale, and general feedback was analyzed using a thematic approach, by manually cleaning, sorting, categorizing, and summarizing data using table matrices to answer questions framed by the key areas of inquiry. Two investigators with expertise in qualitative methods, product development, and global health research were involved in the analytic process. One investigator conducted data quality checks in collaboration with data collectors to ensure completeness and consistency. The second investigator reviewed the full dataset, generated a codebook that incorporated both predefined themes from the interview guides (deductive coding) and emergent themes (inductive coding) and synthesized the findings. Coded themes were reviewed by data collectors in each country (co-authors of this manuscript).

### Ethics

Ethical approval for this study was issued by the Mildmay Uganda Research Ethics Committee (MUREC-2024-359), the Uganda National Council for Science and Technology (HS3928ES), and the University of KwaZulu-Natal Biomedical Research Ethics Committee (6312/2023). Participants provided written informed consent in their preferred local language.

## Results

A total of 48 stakeholders participated in the study, of which 9 were decision-makers (in-depth interview only) and 39 were target end users (simulated use, observation, and in-depth interview) (Table 1).

**Table 1.** Study participant characteristics.

| Type of stakeholder | South Africa<br>(n=20) | Uganda<br>(n=28) | Total<br>(n=48) |
| --- | --- | --- | --- |
| Decision-maker | 3 | 6 | 9 |
| Target end user | 17 | 22 | 39 |
| Health care provider | 6 | 8 | 14 |
| *Nursing officer* | 3 | 4 | 7 |
| *Adherence counselor* | 3 | 4 | 7 |
| Community health worker | 3 | 5 | 8 |
| Caregiver (age range 21–59 years) | 8 | 9 | 17 |
| *of a newborn* | 3 | 3 | 6 |
| *of an infant up to 1 year old* | 3 | 4 | 7 |
| *of a child 2 to 3 years old* | 2 | 2 | 4 |

### Usability and additional feedback

For the target end users who underwent simulated use and observation, the MAP indicator prototype usability metrics (observed completion of steps, participant-reported ease of use and confidence) were all 89% or greater (Table 2). A limited number of use errors and use difficulties were observed (Table 3).

**Table 2.** Usability metrics (n=39).

| Observed | Number of participants<br>(percentage) |
| --- | --- |
| Completed all steps included in job aid during unguided simulation | 38 (97.4%) |
| Used job aid during unguided simulation | 35 (89.7%) |
| Reported by participant | Number of participants<br>(percentage) |
| Felt that using the prototype overall was very easy or easy | 37 (94.9%) |
| Felt very confident or confident they had successfully administered the prototype | 38 (97.4%) |
| Recognized at least two of the three indicator cues (tactile, visual, auditory) as confirmation of successful delivery | 39 (100%) |

**Table 3.** Observed use errors and use difficulties (n=39).

| **Use error** | **Number of participants (percentage)** |
| --- | --- |
| Participant did not press hard enough on dome, so it did not snap and it reinverted | 4 (10.3%) |
| **Use difficulty** | **Number of participants (percentage)** |
| Lid broke off during opening of MAP tray | 2 (5.1%) |
| Despite participant pressing correctly, dome changed color but did not snap, or dome reinverted and had to be pressed again to change color | 7 (17.9%) |
| Skin adhesive and indicator were stuck together, so participant could not easily remove indicator to leave the skin adhesive on manikin skin | 3 (8%) |
| Participant struggled and took a long time to peel off the patch from the manikin skin | 8 (20.5%) |

Even though all participants recognized the tactile and visual cues (feeling the dome invert and seeing the dome change color from transparent to white), three participants did not recognize the auditory cue (hearing the snap upon pressing the dome). The majority of participants (28/39, 71.8%) thought that seeing the dome change color was the best cue to confirm correct delivery. Some participants (6/39, 15.4%) thought that an auditory cue would be easily missed in noisy settings.

Participants shared the following reasons why they felt the prototype was easy to use:

- “Instructions are clear and easy to follow. Anyone can follow them.” —Nursing officer, South Africa
- “Not much work, compared to oral treatment.” —Caregiver, Uganda
- “It was easier than administering oral syrup, which I am giving to my child right now. Sometimes he even vomits the drug, and I have to re-administer.” —Caregiver, Uganda
- “Better than fighting with a child to take oral treatment.” —Caregiver, South Africa

Some use difficulties and use errors were identified during observation of unguided simulation. Table 3 reports on the difficulties and errors observed in at least two or more participants.

Participants who either missed a step or who experienced a use error explained that this was their first time using the MAP, that how they positioned the manikin affected their ability to press the dome, and that they felt nervous they would hurt “the baby” while pressing the dome but that with practice they could learn how to use it correctly. Possible explanations for the observed use difficulties are described in the Discussion section.

When asked if they would change anything about the prototype, the majority of participants (28/39, 71.8%) said they would not change anything:

- “I like it this way. It will be easy to use by older people.” —Adherence counselor, South Africa
- “It should be released for our use immediately.” —Community health worker, Uganda

Those who had suggestions wanted the dome to be easier to press, and peeling the indicator off the (manikin) skin to be easier to do.

### Acceptability and programmatic fit

All 48 participants, including both target end users and decision-makers, said the MAP indicator prototype was acceptable to deliver the needed ARV medication to newborns and young children because it would be easy to administer, discreet, convenient to apply once weekly or monthly, more comfortable than administering daily oral medication, and prevent vomiting and likely improve adherence. Some health care providers also mentioned that it could potentially improve mental health, as children and their caregivers would not have to think about the disease every day, and that it would make the lives of caregivers much easier. Caregivers expressed a sense of urgency to make these MAPs available for use as soon as possible: “When are you giving us these?” —Caregiver, Uganda

Almost all participants thought the MAP size and the expected temporary skin changes (explanation in lay terms: “it could look red, or it could become a bit darker, or little white dots could be visible”) after MAP application would be acceptable (Table 4), given that the MAP is small enough to handle and allows privacy, and because caregivers are already used to temporary skin side effects after immunization.

**Table 4.** Perceptions of acceptability of specific microarray patch attributes.

| Perception | Number of participants<br>(percentage) |
| --- | --- |
| MAP size will be acceptable to other health care providers, community health workers, and caregivers | 45/48 (93.8%) |
| Temporary skin changes after MAP application are acceptable | 45/48 (93.8%) |

The subset of participants who were asked about acceptability of the minimum (20 minutes) and maximum wear times (24 hours) for the MAP thought these times would be acceptable (Table 5) because application would be happening at most once a week and because daily oral administration is so challenging.

**Table 5.** Perceptions of acceptability of microarray patch wear time.

| Perception | Number of participants<br>(percentage) |
| --- | --- |
| 20-minute wear time is acceptable | 7/7 (100%)* |
| 24-hour wear time is acceptable | 15/20 (75%)** |

\*Due to limited participant time availability, this question was asked of only seven participants in Uganda.
\*\*The 24-hour wear time question was asked of only 20 South African participants.

The majority of participants found that applying multiple MAPs at once to achieve the necessary dose for a weight band would be acceptable (Table 6). However, not many participants thought applying MAPs sequentially (applying one or more MAPs first, waiting the 20-minute wear time duration, removing those first MAPs, then applying more MAPs, and completing the rest of the steps) would be an acceptable solution (Table 6) because it would be confusing and potentially lead to errors and underdosing.

**Table 6.**
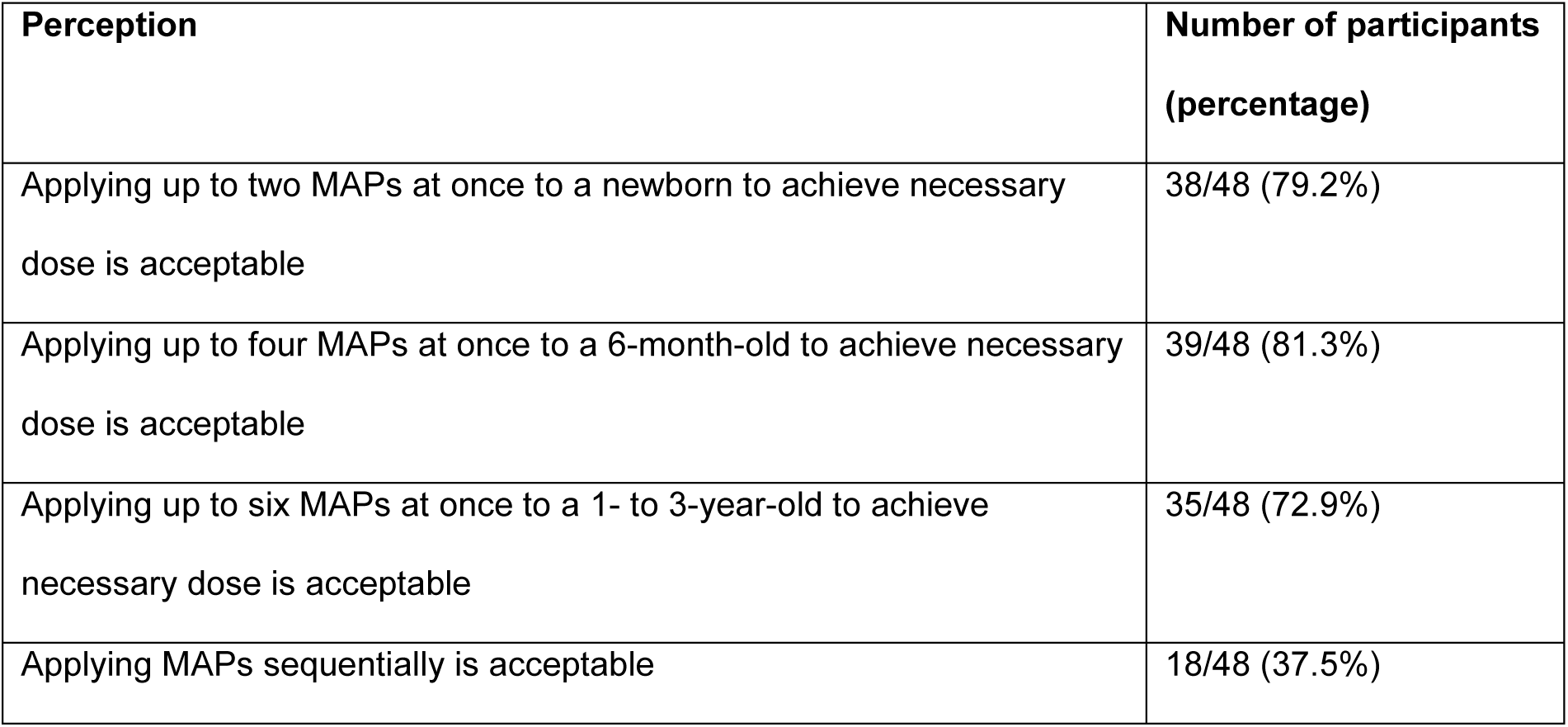
Perceptions of acceptability of microarray patch application modalities.

| Perception | Number of participants<br>(percentage) |
| --- | --- |
| Applying up to two MAPs at once to a newborn to achieve necessary dose is acceptable | 38/48 (79.2%) |
| Applying up to four MAPs at once to a 6-month-old to achieve necessary dose is acceptable | 39/48 (81.3%) |
| Applying up to six MAPs at once to a 1- to 3-year-old to achieve necessary dose is acceptable | 35/48 (72.9%) |
| Applying MAPs sequentially is acceptable | 18/48 (37.5%) |

All participants preferred the possibility of administering a weekly ARV MAP to children than to continue administering daily oral ARVs (Table 7) because oral administration is such a challenge. The majority of participants preferred the possibility of administering a weekly ARV MAP than the possibility of the child receiving an intramuscular ARV injection every 2 to 6 months (Table 7) because children are already receiving many painful immunization injections, MAPs can conveniently be administered at home versus waiting in long queues at health facilities, and the longer administration intervals would not allow for accurate dosing in young children, who change weight bands frequently as they grow.

**Table 7.** Preference for antiretroviral treatment delivery modality.

| Preference | Number of participants<br>(percentage) |
| --- | --- |
| Preference for MAP weekly administration versus daily oral administration | 48/48 (100%) |
| Preference for MAP weekly administration versus an intramuscular injection every 2 to 6 months | 39/48 (81.3%) |

Country decision-makers foresaw that pediatric ARV MAPs could be used for HIV postnatal prophylaxis and maintenance treatment mostly for home delivery by the caregiver, with an initial application at the health facility by the nurse or adherence counselor. They thought that a weekly schedule would be acceptable to health systems if administered by caregivers, given benefits of a MAP over the current standard of care.

## Discussion

This is, to our knowledge, the first study to assess the usability and acceptability of a MAP indicator prototype for pediatric drug delivery in high-burden HIV settings. Our findings suggest that the MAP indicator prototype is highly usable, even among diverse groups of target end users, including health care providers in urban areas and grandmothers caring for children with HIV in rural areas. The majority of participants successfully completed the required steps to administer the prototype on a manikin, felt it was easy to use, expressed confidence in their ability to use it, and recognized at least two cues signaling successful delivery. These results underscore the prototype’s potential to meet the needs of users with varying levels of familiarity with medical devices. Importantly, participants expressed that administering a MAP was easier than providing oral ARVs to children. Many caregivers expressed frustration with the daily challenges of administering oral medication—such as vomiting—which they felt the MAP could alleviate. They found the MAP instructions clear and easy to follow and predicted that it would also be easy for others to follow. This reinforces the potential of the MAP to address key barriers to adherence in pediatric HIV treatment.

Use difficulties and use errors were identified, which is expected in the early stages of prototype testing with target end users and with the limitations inherent in simulating application of a medical-grade adhesive to a plastic baby care model. Some use difficulties, such as the dome reinverting or failing to produce an audible snap upon application of correct pressure, can be attributed to the pre-commercial– scale manufacturing quality of the prototype. We anticipate these issues can be addressed during future iterations by refining both the design and the manufacturing process. Other use difficulties, such as the challenging removal of the adhesive from the manikin skin, are limitations of the manikin and simulated nature of the study. We expect the medical-grade adhesive used in the manufacture of the prototype would be easily peelable from real human skin. This use difficulty will need to be evaluated with prototypes in children.

The use error that was observed in more than one participant (e.g., not pressing hard enough on the dome, causing it to reinvert and not make a snap sound) was attributed by participants to their lack of familiarity with a new device and their nervousness that they could hurt “the baby.” They expected this would change with practice. This suggests that implementation of the pediatric ARV MAP will need to be accompanied by proper training that includes opportunities to practice with prototypes, videos of administration, and supervision of end users during their first administration.

The tactile cue (dome inversion) and visual cue (color change) were universally recognized as successful indicators, while the auditory cue (snap sound) was seen as potentially less reliable, such as in noisy environments. This suggests that future iterations may need to prioritize the tactile and visual cues to ensure broad usability, especially to meet the needs of users with sensory limitations. However, when administered in a quieter home environment, the auditory cue might be more easily appreciated.

The pediatric ARV MAP prototype was overwhelmingly accepted by participants, who viewed it as a promising solution to the palatability and adherence challenges posed by current oral pediatric ARV formulations for lifelong treatment. A sense of urgency to have access to such a solution was conveyed by participants who face this challenge every day. Other studies conducted in low-, middle-, and high-income countries have also reported high acceptability of MAPs for pediatric use by health care providers and caregivers, but within the context of administration of vaccines [19,20,21]. A survey conducted with health care providers and caregivers in Egypt that explored acceptability of MAPs specifically for pediatric drug delivery also found that stakeholders were willing to use them [30]. In contrast, in a study conducted in South Africa, a transdermal patch was the least favored ARV option among mothers of newborns living with HIV and health care providers [31]. The study was nested within a dolutegravir (DTG) dispersible tablet and oral film clinical trial and asked participants to rank their preference for different formulations, This relative inferior perception of a MAP can likely be explained by several differences in study methods: that study only provided participants with images of the hypothetical patch option, used the term “microneedles” to describe it, set a wear time of a couple of days, and conducted a ranking exercise. Our study provided participants with a physical prototype that they applied on a manikin, used the term “microprojections” to describe the patch, set a wear time of 20 minutes (or 24 hours in some cases), and asked about overall and specific acceptability. In addition, mothers in the other study had been using either one of the new oral DTG formulations as part of the study, so familiarity with those formulations could have also impacted their ranking. Our participants explicitly compared their experience of daily administration of an oral syrup with the possibility of a weekly patch which they had now seen and touched.

The compact size of the pediatric ARV MAP prototype, its discreet application, and the potential for a less frequent administration schedule were all seen as advantages. While the possibility of applying multiple MAPs sequentially to meet required dosages raised concerns about complexity and dosing errors, application of multiple MAPs at once was acceptable to achieve the necessary dosage in different age groups. This contrasts with the findings of a study conducted in Ghana, Kenya, and Uganda, where women of reproductive age did not find the requirement of applying multiple contraceptive MAPs to achieve a longer duration of protection acceptable [32]. This difference might be explained by two factors: (1) difference in study methods: participants in the contraceptive MAP study were only shown drawings of the MAPs, so their perception could have been different if they had handled a real-life prototype or if they had engaged in simulated use, like our participants did; (2) difference in use case: caregivers of children living with HIV face challenges with oral administration on a daily basis, with life and death consequences for their children, making a weekly application of multiple patches more acceptable.

In our study, when participants were asked if they preferred administering a weekly MAP, their current daily experience of oral administration, or the possible option of an intramuscular injection every 2 to 6 months, the majority preferred the weekly MAP option. Acceptability of this schedule is both different and similar to what a study [33], also conducted in South Africa and Uganda, found regarding an HIV pre-exposure prophylaxis (PrEP) MAP. Most of the young adults who participated in that study preferred a duration of protection of 1 month or longer, but some preferred a weekly option to match their sexual activity frequency. The difference in schedule preference in our study might be explained again by two factors: (1) difference in study methods: participants in our study were asked about acceptability of the shortest feasible MAP schedule only, since ARV MAP regimens may be driven by drug potency and loading capacity; (2) difference in use case and perception of risk: children living with or exposed to HIV requiring daily treatment versus healthy young adults at high risk of HIV infection. Ultimately, the pharmacokinetic results from preclinical and clinical trials will determine the appropriate number of MAPs and duration of protection, and feedback from users and stakeholders will inform product developers on how to balance these attributes.

The high acceptability of both the 20-minute and 24-hour wear times (even when in a limited sample size) further supports the flexibility of the MAP as a treatment option, suggesting these wear times may be feasible for real-world use, especially when compared to the current daily oral regimen. Similarly, in the HIV PrEP use case, while participants preferred a shorter wear time (30 minutes), they were willing to accept a longer wear time (1 hour) if that correlated to a longer duration of protection [33]. In contrast, with MAPs used to deliver measles and rubella vaccines as part of immunization campaigns, stakeholders preferred a 1-minute wear time over 5 minutes [34]. The administration challenges noted with the 5-minute wear time included needing to hold the child’s body part still and the time the health worker would be required to spend with each child, which would be longer than for current injectable vaccine administration. These administration challenges are alleviated in the pediatric ARV MAP prototype because (1) it would be applied out of reach of the child and the indicator portion of the MAP would be removed, leaving the microprojections in the child’s skin, and (2) by the use case of application at home by a caregiver.

Our study results suggest that temporary skin changes, expected with MAP application, will be acceptable in the target end user population. Similarly, acceptability of temporary skin changes has been reported after real-life application of vaccine and placebo MAPs in adults [35,36,37]. Clinical trials of placebo and measle-rubella vaccine MAPs have assessed acceptability and reactogenicity in children for immunization use [38,39], but a peer-reviewed publication on the acceptability of the skin changes is not available yet.

From a programmatic perspective, decision-makers indicated that a MAP-based treatment model could be feasibly integrated into pediatric HIV care. Importantly, the advantages of a weekly ARV MAP schedule identified by participants, such as allowing frequent dose adjustment in the first year of life, eliminating long queues at health facilities for intramuscular administration, and avoiding hesitancy associated with injections within the context of an expanding infant immunization schedule, positions the MAP as a viable option to overcome access barriers and the emotional burden of current treatment options.

The strengths of our formative assessment include use of qualitative methods that allowed participants to have a better understanding of how the potential product would feel and work, rendering rich contextual insights from participants, and allowing early identification of the necessary iterative refinement for the product as well as implementation barriers. Limitations of our study include the small sample size and purposive sampling that limit generalizability of findings, and potential observer effect on participant responses.

## Conclusions

The pediatric ARV MAP prototype has high acceptability and high usability among caregivers, community health workers, health care providers, and decision-makers in two high-HIV-burden settings, South Africa and Uganda. Our findings suggest the MAP has the potential to revolutionize pediatric HIV treatment by improving adherence, reducing caregiver burden, and enhancing overall treatment outcomes for children. The acceptability of the pediatric ARV MAP was similarly high to that reported in the literature for other use cases (MAPs for contraception, HIV PrEP, or vaccine delivery). However, the preferred characteristics for a pediatric ARV MAP were sometimes different from those other use cases, as expected. In our study, the preferred characteristics expressed by stakeholders seemed to be influenced by the unique use case, criticality of HIV treatment, and challenging current standard of care. Successful integration of a pediatric ARV MAP into health systems will require comprehensive training, ongoing education, and careful consideration of local contexts. Further development and clinical evaluation of this technology is warranted.

## Data Availability

All data produced in the present study are available upon reasonable request to the authors

## Acknowledgments

We thank the decision-makers, health care providers, and caregivers in South Africa and Uganda for their willingness to participate in this study, and we honor them for sharing their invaluable time and insights. We also recognize the essential work of the South Africa National Department of Health and the Uganda Ministry of Health. At PATH, we thank Jill Sherman-Konkle, Abra Greene, Tessa Fielding, Clara Orndorff, Teri Gilleland, and Patrick McKern for their efforts in support of this work. At Africa Health Research Institute, we thank Leora Sewnarain and Tawanda Makusha for their support.

## Supporting information caption

**S1 Appendix**. Script used to explain how MAPs work.

**S2 Appendix**. Job aid with illustrated steps for using the indicator prototype.

