## Supplementary material for "‘We need this now’: Acceptability and usability of a pediatric antiretroviral microarray patch prototype in South Africa and Uganda": S1 Appendix

### Introduction to microarray patches

Microarray patches (MAPs) are devices that allow delivery of drugs or vaccines just under the skin through microprojections. *(Show/explain the image below, using simple terms for layers of the skin.)*

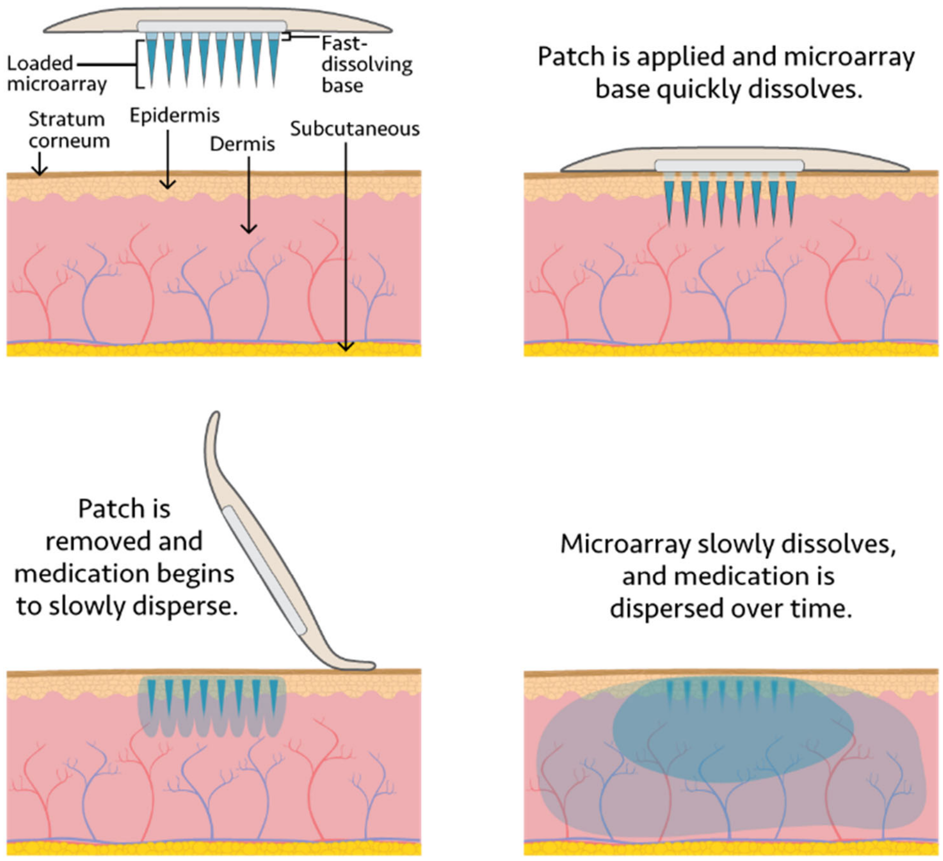


We are developing a MAP containing long-acting HIV medication for children. Based on the latest studies we have done, the MAP will have the following characteristics:

- Depending on the final medication in it, it will need to be applied only once a week or once a month, either to prevent HIV in babies born to HIV-positive mothers or to provide maintenance treatment to HIV-positive young children.
- For babies and young children, the MAP will be administered on the upper back because it's a firm surface and so that the child cannot remove it, so that it doesn't get dirty from the diaper area and so that it can be covered by clothing if the user prefers that.
- Users like you will firmly press the MAP onto the skin to ensure the projections penetrate and dissolve. I will show you in more detail soon.
- This will be perceived by the child as less painful than an injection.
- The MAP will need to stay on the skin for 20 minutes so that the microprojections dissolve and deposit the drug under the skin. During this time, the user can go about their normal activities and the child can nap. When the time is done, the user will remove the MAP and discard it. The drug under the skin will slowly release into the child's body over time.
- The MAP does not require refrigeration (different from some oral HIV medications).
