## Supplementary material for "‘We need this now’: Acceptability and usability of a pediatric antiretroviral microarray patch prototype in South Africa and Uganda": S2 Appendix

### MAP indicator prototype

#### Parts

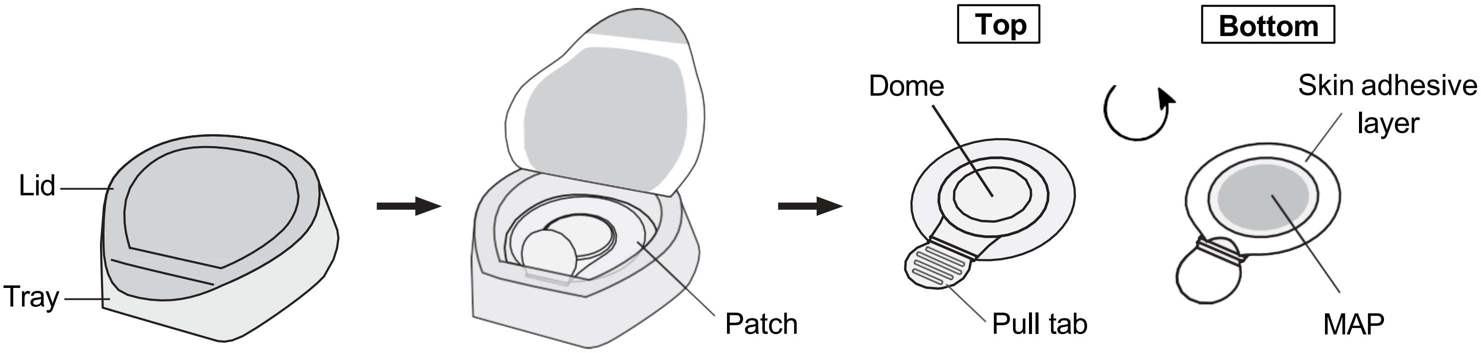


*Note: do not use MAP if indicator dome is crushed*

#### Instructions for use

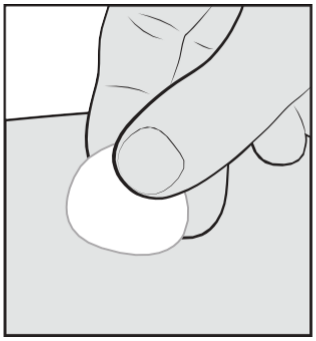

1. Make sure the application site is clean and dry.

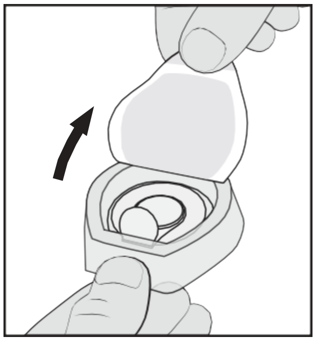

2. Open

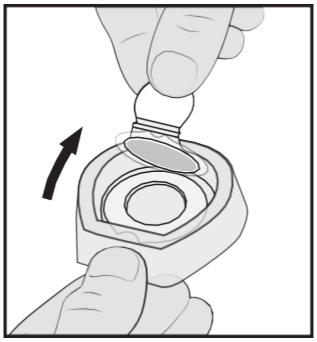

3. Using the pull tab, peel patch out of the tray.

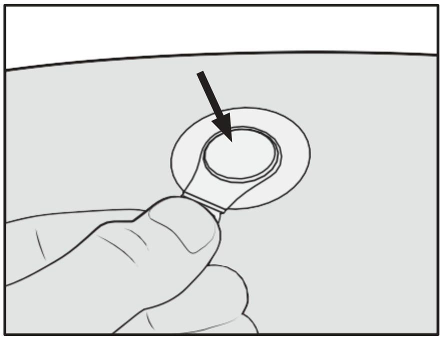

4. Place patch on application site adhesive side down.

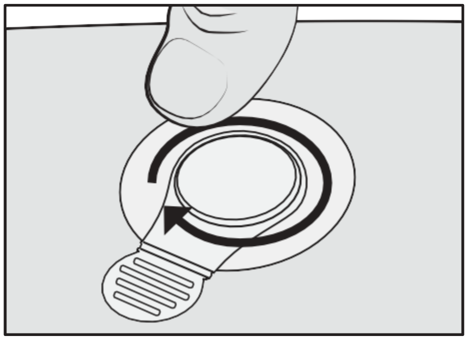

5. Firmly press outer edges of patch so it sticks well to the skin.

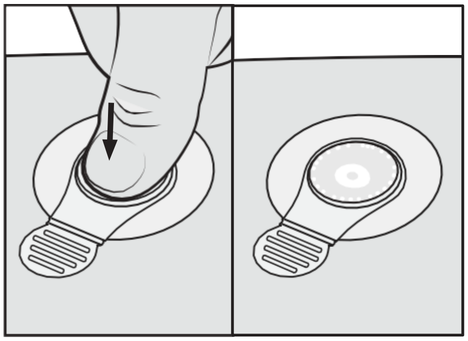

6. Press down firmly on the dome. You should feel the dome crush and hear it snap. The dome should turn white.

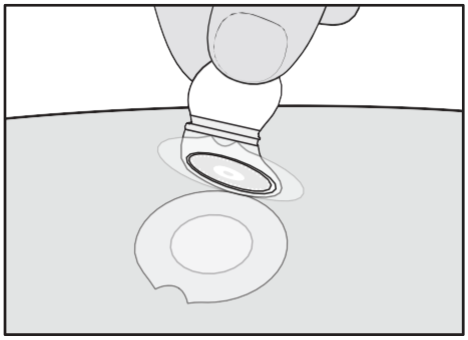

7. Using the pull tab, peel off the plastic dome layer, leaving skin adhesive and MAP on skin.

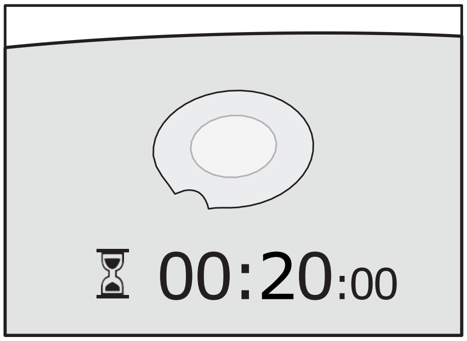


8. Wait 20 minutes.

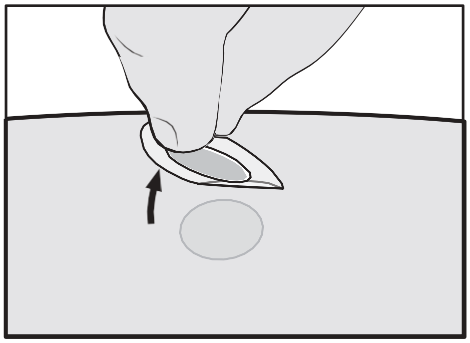

9. Peel patch off application site.

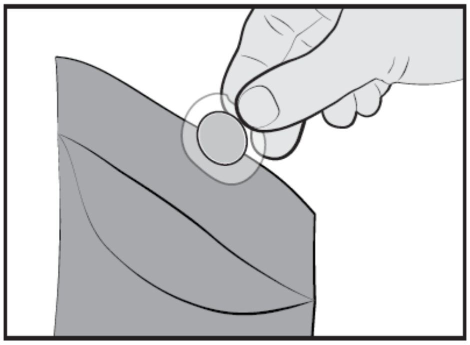

10. Dispose with normal waste.
